# “We are at risk too”: The disparate impacts of the pandemic on younger generations

**DOI:** 10.1101/2020.07.21.20159236

**Authors:** Renée El-Gabalawy, Jordana L. Sommer

## Abstract

**Background:** The COVID-19 pandemic has resulted in profound global impact with high rates of morbidity and mortality. It is essential to understand the psychosocial impacts of the pandemic to identify appropriate prevention and intervention targets. Across generational groups, this study examined: (1) rates of precautions and adaptive and maladaptive health behaviours, (2) differences in levels of anxiety, and (3) rates of and changes in COVID-related concerns over time during the early outbreak of COVID-19 in Canada.

**Methods:** We analyzed data from two Canadian population-based datasets: the Canadian Perspective Survey Series: Impact of COVID-19 survey (*N*=4,627; March 29-April 3, 2020), and Crowdsourcing: Impacts of COVID-19 on Canadians – Your Mental Health (*N*=45,989; April 24-May 11, 2020). We categorized generational age group, participants self-reported changes in behaviours and COVID-related concerns, and a validated measure assessed anxiety symptoms.

**Results:** There are age differences in behavioural responses to the pandemic; adaptive health habits (e.g., exercise) were stable across groups, while maladaptive health habits (e.g., substance use) were highest among younger groups. COVID-related precautions were also highest among the younger generations, with Generation X exhibiting the highest rate of precautionary behaviour. Results also revealed that anxiety and worry are prevalent in response to the pandemic across all generations, with the highest rate of clinically significant anxiety among Millennials (36.0%). Finally, COVID-related concerns are greatest for younger generations and appear to be decreasing with time.

**Conclusion:** These early data are essential in understanding at-risk groups given the unpredictable nature of the pandemic and its potential long-term implications.

## Introduction

The COVID-19 pandemic has resulted in profound global impact. Beyond those acquiring the disease, persons are also affected by physical distancing and additional unprecedented stresses related to financial loss, childcare challenges, and limited healthcare access, among others. It is recognized that aging populations are at elevated risk of adverse outcomes including COVID-related critical illness and death.^1-3^ Less well understood are the psychosocial impacts that will undoubtedly outlive the pandemic; understanding at risk groups is essential.^4,5^ This study provides preliminary insight into COVID-related psychosocial impacts across generational groups in Canada.

Rates of stress and worry are higher among younger generations including “Millennials” and “Generation Xers” compared to older groups including “Baby Boomers” and the “Greatest/Silent Generations”.^6-8^ This transcends to age-related differences in rates of mental disorders, which are elevated in younger compared to older generations.^9^ Charles’ Strength and Vulnerability Integration theory^10^ proposes that older adults have elevated strengths in coping with life stressors and consequently exhibit greater emotional well-being.^6,11^ However, when faced with unavoidable or inescapable negative events, these age-related strengths are compromised. There are also unique challenges inherent to older adults with multimorbidity during the pandemic, including isolation^11^ and loneliness, which may be exacerbated by the “digital divide” (i.e., less virtual access)^5^, and may elevate risk of mental health challenges in this context.

Also unclear are the behavioural generational differences during COVID-19. Generally, engagement in preventative health behaviours improves with increasing age and younger generations are at highest risk of engaging in fewer health-promoting behaviours;^12,13^ this may translate to engagement in poor health behaviours during the pandemic. Stress and anxiety during COVID-19 may also impact behavioural responses; research suggests elevated concern is associated with adherence to behavioural recommendations and adaptive health behaviours.^14,15,16^ However, significantly elevated distress can result in counterintuitive behavioural practices, including maladaptive health behaviours (e.g., increased smoking) or avoidance.^14^ This may suggest that younger generations, who are at potentially higher risk of compromised mental health, may exhibit poorer behavioural responses compared to older adults. In support, research on the SARS outbreak revealed that older adults were better able to adapt their coping strategies to the changing environmental context, and engaged in more precautionary health behaviours.^17,18^

The current study aims to describe behavioural changes, levels of anxiety, and COVID-related concerns across generational groups using two Canadian population-based surveys conducted between March and May 2020. Preliminary research from a convenience sample of 302 adults from the United States revealed that older adults perceive greater risk of COVID-19 than younger adults, but older men were less worried and endorsed the fewest number of behavioural changes.^19^ This preliminary study, however, was limited in its sample size, was demographically homogeneous, and was based in the United States. Across generational groups in Canada, we aim to: (1) report rates of precautions and adaptive and maladaptive health behaviours during the pandemic, (2) assess differences in levels of anxiety, and (3) examine rates of and changes in COVID-related concerns.

## Methods

### Data

We analyzed Statistics Canada Public Use Microdata from two surveys: Canadian Perspective Survey Series (CPSS) 1: Impacts of COVID-19 (*N* = 4,627; collection response rate = 63.9%), and Crowdsourcing: Impacts of COVID-19 on Canadians – Your Mental Health (*N* = 45,989; response rate is unavailable based on sampling method). Data from survey 1 were collected online between March 29-April 3, 2020. The sample is comprised of Canadians aged 15 years and older who live in the 10 provinces; those living in households in extremely remote areas, institutionalized individuals, and those living on reserves were excluded from participating. This survey utilized Statistics Canada’s pilot probability panel as a sampling frame, which was created using a subset of Labour Force Survey (LFS) respondents; the LFS utilizes stratified, multi-stage, probability sampling. Statistical weights were derived to ensure accurate representation of the Canadian population. Additional details are published elsewhere.^20^

Data from survey 2 were collected online between April 24-May 11, 2020. Open advertising recruited participants, and the sample was self-selected. The target population included Canadians, aged 15 years and older, living in any of the 13 provinces and territories. Benchmarking techniques were applied to correct for unbalanced responding across sociodemographics. Additional details are published elsewhere.^21^

### Measures

#### Sociodemographics

We derived generation groups from pre-categorizations by Statistics Canada that approximate definitions from prior research:^22-25^ “Millennials” (ages 15-34), “Generation X” (ages 35-54), “Baby Boomers” (ages 55-74), and the “Greatest/Silent Generations” (ages 75+). Within the survey 2 sample, Baby Boomers were collapsed with the Greatest/Silent Generations due to the highest pre-determined age category being 55+.

Additional sociodemographics are shown in Table 1.

**Table 1.**
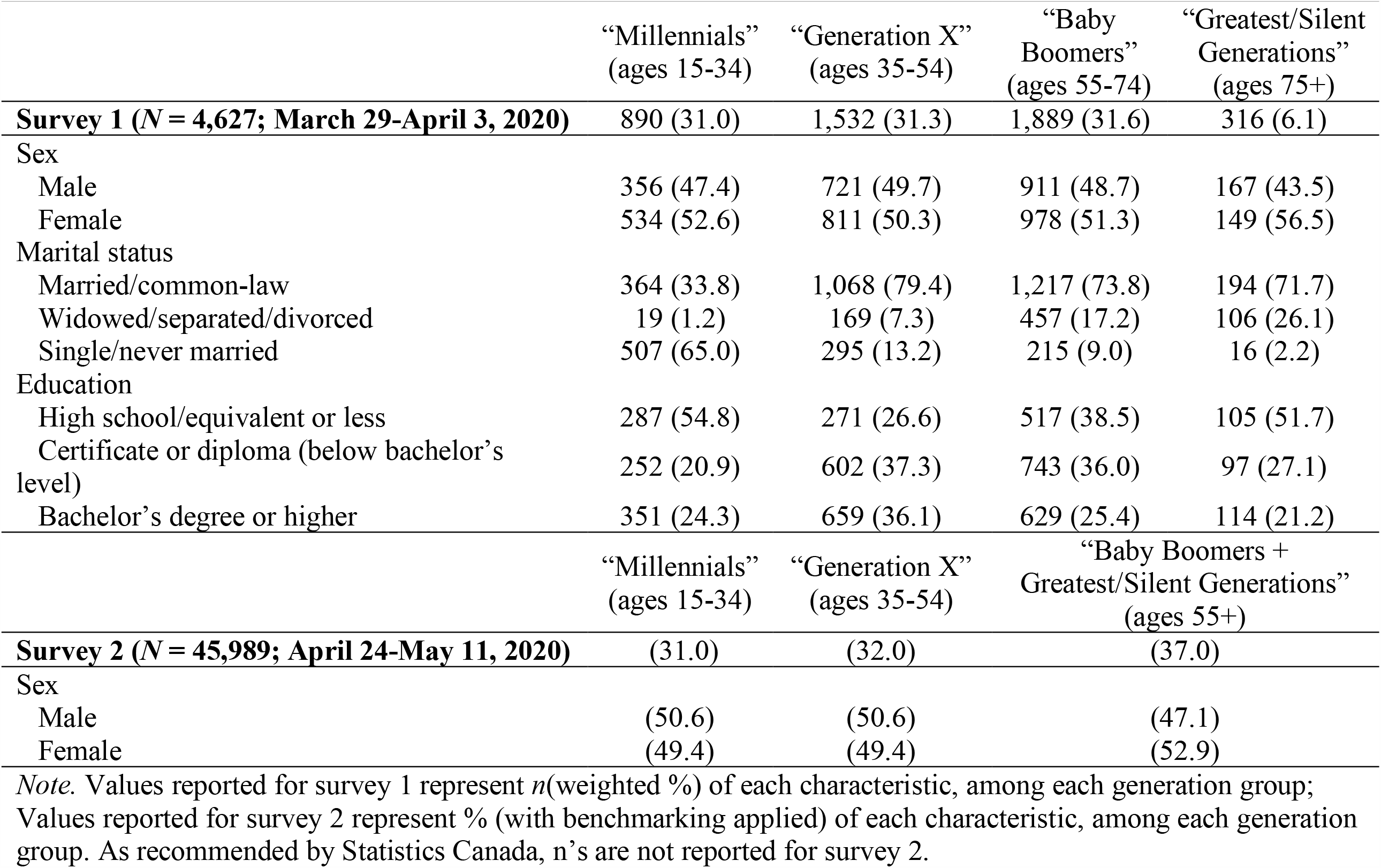
Sample Characteristics According to Generation

#### Behavioural impacts of COVID-19 (survey 1)

Within survey 1, participants were asked, “*Which of the following precautions have you taken to reduce your risk of exposure to COVID-19?*” (*Yes*/*No* for each precaution). Participants were also asked, “*Are you doing any of the following activities for your health?*” (*Yes, for my mental health, Yes, for my physical health, Yes, for both my mental and physical health, No* for each activity). We collapsed all “*Yes*” options to create dichotomized variables for each activity (yes, no). Finally, participants were asked, “*Have your weekly habits changed for any of the following activities?*” (*Increased, Decreased, No change* for each activity); we report on habits that have increased. We also computed a continuous variable, representing the number of behavioural impacts of each kind. See Table 2 for additional behavioural impacts.

**Table 2.** Behavioural Impacts of COVID-19 According to Generation (Survey 1 Only)

|  | “Millennials”<br>(ages 15-34) | “Generation X”<br>(ages 35-54) | “Baby<br>Boomers”<br>(ages 55-74) | “Greatest/Silent<br>Generations”<br>(ages 75+) | *F*-statistic |
| --- | --- | --- | --- | --- | --- |
| **Survey 1 (*N* = 4,627; March 29-April 3, 2020)** | 890 (31.0) | 1,532 (31.3) | 1,889 (31.6) | 316 (6.1) |  |
| **Precautions taken to reduce risk** |  |  |  |  |  |
| Stocked up on essentials | 570 (62.2) | 1,030 (65.8) | 1,218 (62.4) | 199 (54.9) |  |
| Filled prescriptions | 198 (19.2) | 472 (30.1) | 840 (43.4) | 172 (46.6) |  |
| Made plan to care for household members who are ill | 88 (8.1) | 201 (13.4) | 160 (9.1) | 15 (3.6) |  |
| Made plan for non-household members | 139 (17.7) | 331 (20.5) | 307 (17.0) | 15 (4.5) |  |
| Made plan to communicate with family/friends/neighbors | 390 (43.3) | 648 (41.4) | 877 (45.9) | 144 (44.5) |  |
| Avoided leaving the house for non-essential reasons | 805 (90.1) | 1,402 (91.8) | 1,718 (90.1) | 292 (85.3) |  |
| Social distancing in public | 798 (88.9) | 1,370 (89.0) | 1,685 (85.9) | 251 (70.4) |  |
| Avoided crowds/large gatherings | 803 (89.2) | 1,340 (88.0) | 1,671 (86.5) | 270 (76.4) |  |
| Washed hands more regularly | 819 (91.0) | 1,416 (92.7) | 1,768 (92.6) | 295 (90.9) |  |
| Avoided touching your face | 603 (64.8) | 1,085 (72.9) | 1,337 (73.4) | 199 (61.5) |  |
| Cancelled travel | 347 (40.3) | 581 (37.4) | 739 (36.2) | 109 (28.7) |  |
| Worked from home | 342 (35.1) | 646 (38.9) | 370 (18.1) | 18 (4.1) |  |
| Mean number of precautions taken (*SE*) | 6.63 (0.00)<sup>bd</sup> | 6.87 (0.00)<sup>ad</sup> | 6.73 (0.00)<sup>d</sup> | 6.28 (0.00)<sup>bc</sup> | 7.02*** |
| **Adaptive health habits** |  |  |  |  |  |
| Engaged in communication with friends and family | 824 (93.5) | 1,406 (91.6) | 1,706 (91.7) | 290 (92.4) |  |
| Engaged in meditation | 224 (25.7) | 393 (26.4) | 449 (26.9) | 84 (26.4) |  |
| Engaged in exercise outdoors | 542 (59.8) | 1,021 (60.5) | 1,340 (69.4) | 199 (62.8) |  |
| Engaged in exercise indoors | 522 (64.1) | 882 (58.7) | 1,018 (57.0) | 200 (68.7) |  |
| Changed my food choices | 363 (41.4) | 585 (39.2) | 577 (33.1) | 82 (32.1) |  |
| Mean number of current activities for health (*SE*) | 2.78 (0.00) | 2.81 (0.00) | 2.75 (0.00) | 2.79 (0.00) | 0.66 |
| **Maladaptive health habits** |  |  |  |  |  |
| Increased alcohol consumption | 196 (18.7) | 265 (18.5) | 114 (6.5) | 9 (2.9) |  |
| Increased use of tobacco | 41 (3.5) | 55 (4.8) | 39 (2.0) | -- |  |
| Increased cannabis consumption | 96 (11.6) | 72 (6.5) | 33 (1.9) | -- |  |
| Increased junk food consumption | 377 (43.3) | 436 (30.0) | 277 (14.0) | 14 (2.2) |  |
| Watched more television | 581 (68.6) | 904 (65.0) | 1,114 (59.8) | 171 (56.0) |  |
| Spent more time on the internet | 688 (80.3) | 1,037 (68.6) | 1,121 (60.1) | 170 (48.9) |  |
| Played more video games | 353 (47.1) | 272 (18.4) | 138 (7.5) | 19 (5.5) |  |
| Played more board games | 290 (34.5) | 520 (34.3) | 179 (10.2) | 30 (11.9) |  |
| Mean number of weekly habits that have increased (*SE*) | 2.53 (0.00)<sup>bcd</sup> | 2.07 (0.00)<sup>acd</sup> | 1.47 (0.00)<sup>ab</sup> | 1.27(0.00)<sup>ab</sup> | 168.42*** |

*Note.* Values represent *n*(weighted %) of each behavioural impact, among each generation group. -- = cell < 5; \*\*\**p* < .001; <sup>a</sup>Significantly differs from Millennials, *p* < .05; <sup>b</sup>Significantly differs from Generation X, *p* < .05; <sup>c</sup>Significantly differs from Baby Boomers, *p* < .05; <sup>d</sup>Significantly differs from Greatest/Silent Generations, *p* < .05.

#### Mental Health Impacts of COVID-19 (survey 2)

Within survey 2, the 7-item Generalized Anxiety Disorder Scale (GAD-7)^26^ assessed symptoms of anxiety over the past two weeks. The GAD-7 is a valid and reliable self-report measure of generalized anxiety symptoms.^26,27^ A score of ≥10 indicates clinically significant anxiety^28^.

#### COVID-19-Related Concerns (surveys 1 & 2)

Participants from both surveys were asked “*How concerned are you about each of the following impacts of COVID-19?*” (*Not at all, Somewhat, Very, Extremely* for each concern). Only rates of concerns rated “*Very*” or “*Extremely*” are reported. We also computed continuous variables within each survey, representing the number of COVID-related concerns. See Table 2 for the list of concerns.

### Analytic Strategy

First, we assessed the prevalence of each generation, and cross-tabulations assessed sample characteristics according to generation, within each sample. Next, cross-tabulations assessed the prevalence of behavioural impacts of COVID-19 according to generation (survey 1), and analyses of variance (ANOVAs) assessed mean differences in the number of behavioural impacts within each domain (i.e., precautions taken, adaptive health habits, maladaptive health habits), according to generation. A cross-tabulation with a chi-square analysis then assessed differences in the prevalence of clinically significant anxiety according to generation (survey 2). Finally, cross-tabulations assessed rates of COVID-related concerns according to generation, among both samples, and ANOVAs examined whether there were mean differences in the number of COVID-related concerns according to generation. For each set of ANOVAs where the omnibus test was significant, Tukey’s Honest Significant Difference test assessed pairwise comparisons.

For survey 1 data we employed statistical weighting for all reported proportions and applied the adjustment factor (derived by Statistics Canada) to measures of precision to ensure representativeness of the Canadian population. For survey 2 we employed benchmarking for all estimates to correct for unbalanced responding across sex, age group, and province/territory compared to the Canadian population; in line with Statistics Canada’s recommendations, no count totals were reported for these data.^21^

## Results

### Sample Characteristics

Among the survey 1 sample (*N* = 4,627), 31.0% of participants were Millennials, 31.3% Generation Xers, 31.6% were Baby Boomers, and 6.1% were members of the Greatest/Silent Generations. Among the survey 2 sample (*N* = 45,989), 31.0% of participants were Millennials, 32.0% were Generation Xers, and 37.0% were Baby Boomers/Greatest/Silent Generations. See Table 1 for additional sample characteristics.

### Behavioural Impacts of COVID-19 (survey 1)

Table 2 outlines behavioural impacts according to generation. Across generations, over 3/4 reported that they avoided leaving the house, avoided large crowds/gatherings, and washed their hands more regularly to reduce the risk of exposure. Approximately 2/3 of Millennials, Generation Xers, and Baby Boomers reported stocking up on essentials to reduce risk compared to half of members of the Greatest/Silent Generations. Generation Xers more commonly made a plan to care for household members who are ill, compared to other generations (13.4% vs. 3.6-9.1%). There were significant differences in the mean number of precautions taken according to generation (*F*(3) = 7.02, *p* < .001); Generation Xers engaged in the most precautions.

There was similar engagement in all adaptive health habits across generations. Over 90% of participants were communicating with their friends and family, and nearly 60% or more reported exercising. No group differences emerged in the mean number of adaptive health habits.

With respect to maladaptive health habits, Millennials and Generation Xers endorsed the highest rates of increased consumption of alcohol, tobacco, cannabis, and junk food compared to other generations. There were significant differences in the mean number of maladaptive health habits according to generation (*F*(3) = 168.42, *p* < .001), with a linear trend of fewer maladaptive habits with increasing age.

### Mental Health Impacts of COVID-19 (survey 2)

As shown in Figure 2, clinically significant anxiety symptoms were most common among Millennials (36.0%), followed by Generation Xers (27.1%), and Baby Boomers/Greatest/Silent Generations (14.5%). There was a significant difference in clinically significant anxiety according to generation (*χ*^2^(2) = 1,902.25, *p* < .001).

### COVID-19-Related Concerns (surveys 1 & 2; table 3)

Within survey 1, members of the Greatest/Silent Generations expressed greater concerns about their own health (51.5%), the health of their household (61.1%), and the health of Canadians (78.8%) compared to other generations. Baby Boomers expressed greater concerns about violence in the home (9.3%) compared to other generations, and Millennials expressed greater concerns about vulnerable peoples’ health (87.5%) and civil disorder (49.1%). There were significant differences in the mean number of concerns according to generation (*F*(3) = 4.88, *p* < .01); post-hoc analyses revealed that Generation Xers (5.83) reported a greater number of concerns compared to Baby Boomers (5.44).

**Table 3.** COVID-19-Related Concerns According to Generation

|  | “Millennials”<br>(ages 15-34) | “Generation X”<br>(ages 35-54) | “Baby Boomers”<br>(ages 55-74) | “Greatest/Silent Generations”<br>(ages 75+) | *F*-statistic |
| --- | --- | --- | --- | --- | --- |
| **Survey 1 (*N* = 4,627; March 29-April 3, 2020)** | 890 (31.0) | 1,532 (31.3) | 1,889 (31.6) | 316 (6.1) |  |
| *Very or extremely concerned about COVID-19’s impact on...* |  |  |  |  |  |
| My own health | 220 (26.1) | 531 (38.4) | 743 (41.6) | 146 (51.5) |  |
| Health of members of my household | 431 (55.5) | 799 (58.5) | 833 (50.4) | 147 (61.1) |  |
| Vulnerable people’s health | 758 (87.5) | 1,251 (83.2) | 1,342 (74.3) | 166 (64.8) |  |
| Canadian population’s health | 569 (64.6) | 1,049 (71.3) | 1,327 (72.4) | 229 (78.8) |  |
| World population’s health | 561 (67.0) | 1,021 (69.0) | 1,339 (72.7) | 228 (72.7) |  |
| Overloading the health system | 758 (86.5) | 1,291 (84.7) | 1,560 (82.5) | 260 (81.6) |  |
| Civil disorder | 402 (49.1) | 633 (46.1) | 553 (31.1) | 87 (29.9) |  |
| Maintaining social ties | 294 (35.9) | 465 (35.1) | 563 (30.7) | 99 (32.5) |  |
| Ability to cooperate/support one another *during* the crisis | 372 (42.3) | 627 (45.5) | 651 (37.9) | 121 (44.6) |  |
| Ability to cooperate/support one another *after* the crisis | 346 (40.5) | 600 (43.8) | 594 (35.1) | 102 (32.6) |  |
| Family stress from confinement | 308 (36.8) | 517 (37.7) | 454 (24.3) | 61 (23.4) |  |
| Violence in the home | 76 (8.1) | 117 (7.5) | 138 (9.3) | 14 (5.4) |  |
| Mean number of COVID-19-related concerns (*SE*) | 5.73 (0.00) | 5.83 (0.00)<sup>c</sup> | 5.44 (0.00)<sup>b</sup> | 5.47 (0.00) | 4.88** |
|  | “Millennials”<br>(ages 15-34) | “Generation X”<br>(ages 35-54) | “Baby Boomers + Greatest/Silent Generations” (ages 55+) |  | *F*-statistic |
| **Survey 2 (*N* = 45,989; April 24-May 11, 2020)** | (31.0) | (32.0) | (37.0) |  |  |
| *Very or extremely concerned about COVID-19’s impact on...* |  |  |  |  |  |
| My own health | (21.0) | (25.7) | (27.3) |  |  |
| Health of members of my household | (44.4) | (42.0) | (35.0) |  |  |
| Vulnerable people’s health | (78.1) | (75.4) | (61.1) |  |  |
| Canadian population’s health | (50.9) | (46.5) | (45.5) |  |  |
| World population’s health | (51.0) | (45.8) | (49.6) |  |  |
| Overloading the health system | (63.2) | (58.4) | (53.4) |  |  |
| Civil disorder | (34.0) | (27.8) | (21.2) |  |  |
| Maintaining social ties | (39.0) | (33.5) | (29.5) |  |  |
| Ability to cooperate/support one another *during* the crisis | (38.1) | (32.4) | (25.1) |  |  |
| Ability to cooperate/support one another *after* the crisis | (41.0) | (34.2) | (23.4) |  |  |
| Family stress from confinement | (35.6) | (32.7) | (18.2) |  |  |
| Violence in the home | (2.7) | (1.3) | (0.6) |  |  |
| Mean number of COVID-19-related concerns (*SE*) | 4.99 (0.02)<sup>bc</sup> | 4.55 (0.02)<sup>ac</sup> | 3.88 (0.02)<sup>ab</sup> |  | 577.54*** |

*Note.* Values reported for survey 1 represent *n*(weighted %) of each COVID-19-related concern, among each generation group; Values reported for survey 2 represent % (with benchmarking applied) of each COVID-19-related concern, among each generation group. \*\**p* < .01, \*\*\**p* < .001;
<sup>a</sup>Significantly differs from Millennials, *p* < .05; <sup>b</sup>Significantly differs from Generation X, *p* < .05; <sup>c</sup>Significantly differs from Baby Boomers (and Greatest/Silent Generations for survey 2), *p* < .05; <sup>d</sup>Significantly differs from Greatest/Silent Generations (for survey 1), *p* < .05.

**Figure 1.**
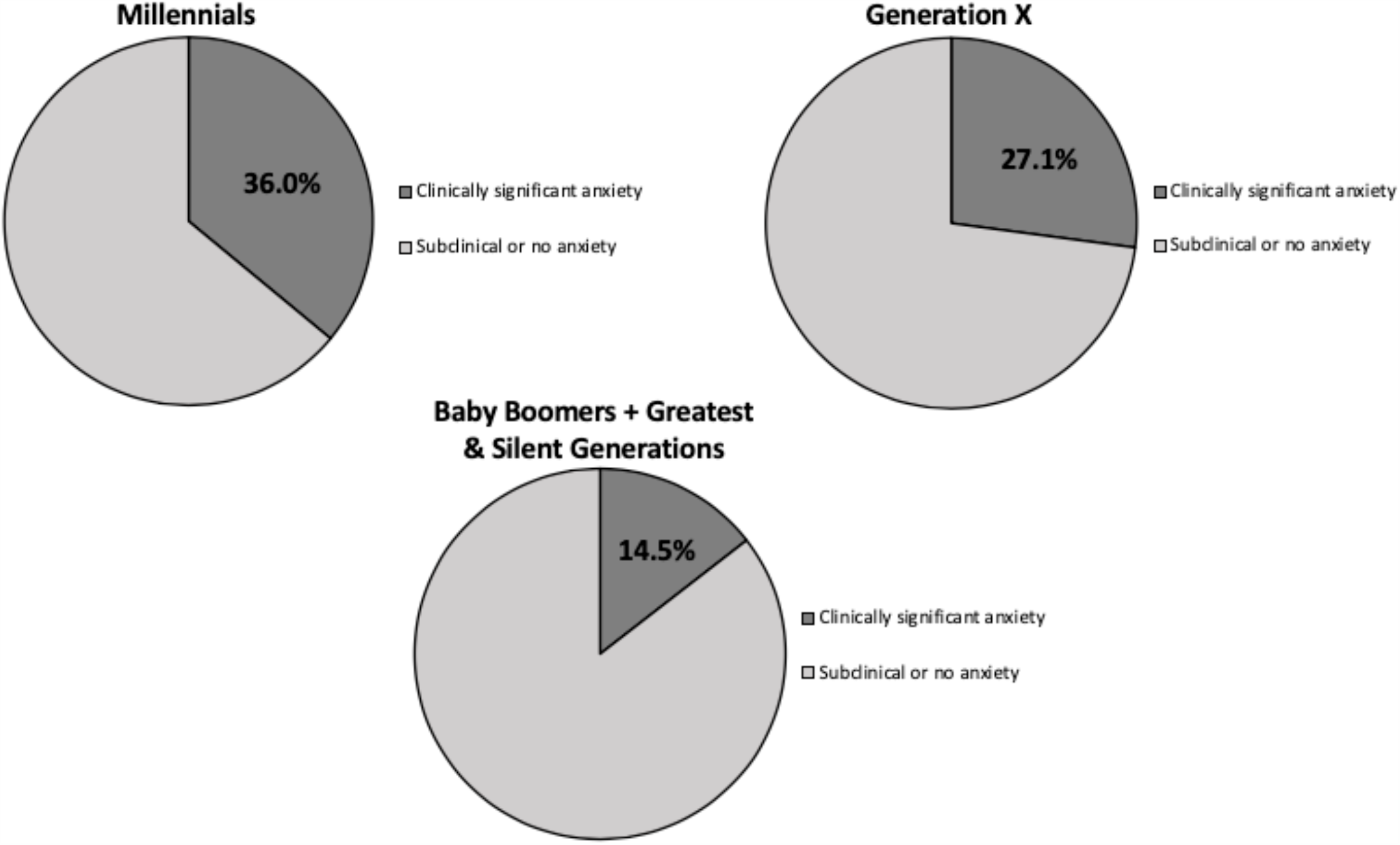
Clinically significant anxiety symptoms according to generation (survey 2: N = 45,989; April 24-May 11, 2020)

Within survey 2, generational trends were maintained but concerns were less common across the majority of domains for each generation, with a few exceptions. For example, concern about maintaining social ties were more common for Millennials and remained similar for Generation Xers and Baby Boomers/Greatest/Silent Generation. There were significant differences in mean number of concerns according to generation (*F*(2) = 577.54, *p* < .001); results demonstrated a linear trend with a decreasing number of concerns according to increasing age.

## Discussion

Given the unprecedented nature of the COVID-19 pandemic, limited information exists on disparities in impacts across the lifespan. Popular media has referenced differential responses across generations but empirical evidence is lacking. This study represents the first population-based examination of behavioural, cognitive, and emotional responses to COVID-19 across generational groups, using population-based Canadian survey data. Three primary results emerged: 1) overall anxiety and worry are prevalent in response to the pandemic across all generations, 2) there are generational differences in emotional and behavioural responses, and 3) COVID-related concerns appear to be decreasing with time. Although COVID-related worries and anxiety are greatest for younger generations, adaptive health habits were stable across groups, while maladaptive health habits were highest among younger groups. Despite this, COVID-related precautions were also highest among the younger generations, with Generation X exhibiting the highest rate of precautionary behaviour.

Mental health trends were consistent with prior research demonstrating that the youngest generation had the highest rate of clinically significant anxiety (36.0%), followed by Generation X (27.1%), and Baby Boomers/Greatest/Silent Generations (14.5%). Comparably, in prior Canadian research using the GAD-7, only 6.1% of adults ages 65+,^29^ 18.6% of a large sample of public safety personnel,^30^ and 15.5% of adults in the Quebec population^31^ scored in the clinically significant level. This suggests that current rates of anxiety in response to COVID-19 are substantially higher than the norm. Disparate trends emerged for specific COVID-related worries; younger generations had a higher mean number of worries relative to older generations. However, the *types* of worries differed across groups. While older generations were more concerned about health and well-being of themselves, their household, and Canadians in general; younger generations had greater concerns about secondary implications of COVID-19 such as civil disorder, maintaining social ties, family stress, and the ability to cooperate. The trend of overall reduced impact of COVID-19 on older generations’ mental health relative to younger generations is consistent with prior trauma research.^32^ This is also partially consistent with one recent COVID-19 study that found that older men reported less worry related to COVID-19 compared to their younger counterparts.^19^ The reduced mental health impact may be related to adaptive coping and emotional regulation strategies apparent in late life.^10^

There were also pandemic-related generational differences in behavioural responses.

Younger generations engaged in a greater number of precautionary behaviours, with Generation X exhibiting the highest level of precautions. Elevated precautions among this group may relate to elevated levels of worry and anxiety^33^ compounded by increased responsibilities in caring for both children and older adults. This is exemplified by the elevated rate of “making a plan to care for household members who are ill” (13.4%) and “concern about family stress from confinement” (37.7%) among Generation X compared to other groups. This is also consistent with Barber and Kim who found lower rates of engagement in precautions among older adults.^19^

Despite these differences, all groups demonstrated similar levels of adaptive health habits. Conversely, a significant proportion of individuals reported changing weekly habits, many of which would be considered maladaptive, and rates differed across groups. Millennials and Generation X reported increased rates of cannabis and alcohol consumption. This may relate to elevated anxiety symptoms among these younger groups, which is recognized to elicit maladaptive health behaviours in general^14^ and in the context of trauma.^34,35^ These behaviours should be closely monitored as they may increase the risk for substance use disorders or other health sequelae.^36^

The two surveys, administered 1 month apart, demonstrate that worries across all age groups appear to decline over time, particularly among older generations. The earlier survey was employed at the early stages of the pandemic in Canada and may have captured acute and elevated concerns related to pervasive uncertainty at that time. Mass media began to display 24/7 coverage of COVID-19, which has been shown to elicit widespread stress in the context of community trauma.^37^ However, with time, a significant proportion of people likely habituate and adjust to these changes and emotional responses, which has been previously described.^38^ Prior research has also shown that older adults tend to exhibit greater responsiveness and flexibility in coping strategies over the course of pandemics,^17^ which may be reflected by larger declines in worries among older groups.

Despite the strengths of the current study including a large population-based sample of contemporary and timely data, there are limitations. First, these trends are based on self-report as opposed to clinical assessments/semi-structured interviews. Second, given time constraints, Statistics Canada was unable to employ rigorous sampling procedures (for survey 2) which may limit generalizability. Third, both surveys were cross-sectional and therefore longitudinal trends could not be examined. Fourth, this study utilized public access data and therefore groups were aggregated to protect the identity of participants; therefore, there were limitations in how we defined generational groups. Finally, findings were based on Canadian data and trends may not be representative of other countries.

The COVID-19 pandemic has resulted in widespread fear and anxiety as indicated by the elevated mental health rates across all generational groups. Clinically significant anxiety ranged from 14.5-36.0%, with Millennials exhibiting the highest levels of both worries and anxiety.

Trends in both precautions and maladaptive behaviours varied according to generation. While members of the Greatest/Silent Generations engaged in the fewest precautionary behaviours, Millennials endorsed the highest number of maladaptive behaviours; interestingly, all generations engaged in similar rates of adaptive health behaviours. One distinction that will be important to further investigate is differentiating adaptive precautions from maladaptive safety seeking behaviours (e.g., excessive precautions in the absence of objective risk) that serve to maintain anxiety long-term.^39^ Mental health should be carefully monitored across the lifespan, particularly younger adults, as early acute stress responses are predictors for adverse health sequelae.^40^ This is particularly problematic given that younger generations are also coping with greater maladaptive behaviours (e.g., elevated alcohol consumption), further exacerbating risk. Finally, targeted interventions such as media campaigns^16^ should be developed to facilitate precautionary behaviours among older populations with the likelihood that active cases will wax and wane over time.

## Data Availability

The analysis is based on public use data that can be accessed through Statistics Canada.

## Acknowledgements

The authors acknowledge Ms. Gabrielle Logan for her contribution of a literature search for the current study. Dr. El-Gabalawy would also like to acknowledge Ms. Bernice Parent, who provided childcare during COVID-19 allowing this research to be made possible. Finally, Dr. El-Gabalawy is funded by University of Manitoba Start-Up Funding and Ms. Sommer receives doctoral funding through the Canadian Institutes for Health Research.

